# Undernutrition and associated factors among lactating mothers in North Shewa Zone, Amhara Region, Ethiopia, 2023. Community based cross-sectional study

**DOI:** 10.1101/2025.10.10.25337752

**Authors:** Mikyas Arega Muluneh, Michael Amera Tizazu, Fetene Kassahun Amogne

**Affiliations:** Department of Midwifery, Health Scince College, Debre Markos University; Department of Public Health, Asrat Woldeyes Health Scince Campus, Debre Berhan University; Department of Midwifery, Asrat Woldeyes Health Scince Campus, Debre Markos University

**Keywords:** Undernutrition, Lactating mother, North Shewa, Ethiopia

## Abstract

**Background:** Health and nutritional status of women are closely linked with the overall health and nutrition of the population. The nutritional demands of lactating women are greater than at any other stage of a women’s reproductive life. Undernutrition during lactation can lead to poor cognitive development in children, reduced immunity, growth faltering, reduced quality and quantity and quantity of breast milk, and increased morbidity and mortality for both in mother and child.

**Objective:** To assess the magnitude of undernutrition and associated factors among lactating mothers North Shewa Zone, Amhara Region, Ethiopia, 2023.

**Method:** Community based cross-sectional study was conducted on 722 lactating mothers with children aged 6-23 months in North Shewa Zone, Ethiopia, from January 01-30/2023. Data were collected using a semi-structured questionnaire. Nutritional status was assessed using Body mass index (BMI). Simple random sampling was used to select study participants. Data were entered into Epi data version 3.1 and exported to SPSS version 25 for further analysis. Bi-variable logistic regression analysis was conducted to identify potential covariates. Variables with a p-value < 0.2 in the bivariate analysis were into a multi-variable logistic regression model to identify independent predictors of undernutrition, with a significance set at P-value < 0.05.

**Result:** Anthropometric measurements showed that the women had a mean BMI of 22.9 ±3.5 kg/m^2^. Among the respondents, 450(66.4%) were of normal weight, 42(6.3%) were obese, 126(18.7%) were over-weight, and 58(8.6%) were under-weight. Factors associated with undernutrition among lactating mothers in this study included being housewives, having a family size five or more, breastfeeding for more than 12 months, lack of postnatal follow up and not using contraceptives.

**Conclusion:** Two-thirds of lactating mothers in North Shewa Zone were within the normal BMI range (18.5 – 24.9kg/m^2^). The prevalence of underweight among lactating mothers was relatively low when comparing with similar communities in Ethiopia. Based on the findings, it is recommended to promote family planning to limit household size and to provide targeted nutritional support for mothers who breastfeed for extended periods.

## Introduction

Malnutrition in all its forms includes undernutrition, overnutrition, and micronutrient deficiencies, each of which impairs physical and cognitive functioning across the lifespan(1). Undernutrition often signaled by low body mass index (BMI < 18.5 kg/m^2^)—is particularly prevalent in low-resource settings and poses significant health risks(1, 2). Nutrition remains the cornerstone of human health, enabling growth, productivity, and resilience through life’s stages(2). Women’s nutritional needs surge during pregnancy and lactation, making these periods especially critical for targeted nutritional support(2, 3).

Nutrition is foundational to sustainable development: it supports health, education, gender equity, poverty reduction, and overall human potential(3). Access to adequate, nutritious food is recognized as a basic human right(4). The health status of women strongly influences that of their families and communities, particularly through maternal and child health outcomes(5). The nutritional quality during the first 1,000 days from conception until a child’s second birthday— determines lifelong physical and cognitive potential(6).

Lactation imposes the highest nutritional demands on women during their reproductive phase. Mothers producing 700–800 mL of breast milk daily require roughly an additional 500-640 kcal/day, alongside increased micronutrient intake (6, 7). When dietary intake is inadequate, maternal nutrient stores deplete, leading to diminished breast milk quality and quantity, which adversely impacts both maternal and infant health(7, 8).

Globally, over 1 billion women aged 15–49 suffer from one or more micronutrient deficiencies especially in regions across sub-Saharan Africa and South Asia, where roughly 10% of women are underweight and about 30% have anemia(1). Moreover, Africa bears more than one in five of the global chronic undernourishment burdens, with economic consequences including estimated losses of 11% of GDP annually(9).

Maternal undernutrition is linked to increased morbidity and mortality-for example, anemia contributes to up to 20% of maternal mortality, while inadequate nutrition in pregnancy increases susceptibility to obstetric complications(8, 10). Undernourished mothers are more likely to give birth to low birthweight infants, contributing to around 3 million maternal and child deaths annually and affecting 45% of under-five mortality due to malnutrition(11).

Child and maternal undernutrition continue to pose serious public health challenges across the globe. Approximately 90% of stunted children live in Africa and Asia, and millions of adolescent mothers begin lactation in developing countries each year, putting both mother and child at risk(11, 12). In Ethiopia, national surveys show that over 22–25% of women of reproductive age are underweight(13). The prevalence among lactating mothers varies widely from around 17% in Arba Minch to up to 40% in some districts(7, 14, 15).

Recent community-based studies across Ethiopia report undernutrition prevalence in lactating women ranging from 19% to 27% in areas like Yilmana Densa, Dire Dawa, Chiro, and Angecha(3, 6, 15, 16). In humanitarian settings like Tigray and Sekota IDP camps, the prevalence is even higher (38%) when assessed via MUAC(4, 17). Associated factors consistently include low dietary diversity, household food insecurity, low maternal education, large family size, inadequate antenatal care, and limited infrastructure such as latrine availability(3, 4, 6, 8, 16, 17).

Despite the burden, evidence remains fragmented particularly for regions like North Shewa Zone. Understanding the magnitude and determinants of undernutrition in lactating women is essential to tailor interventions, inform policy, and break the intergenerational cycle of malnutrition. This study aims to assessed the prevalence of undernutrition and identify associated factors among lactating mothers in North Shewa Zone, Ethiopia.

## Methods and Materials

### Study design, area and period

A community-based cross-sectional study was conducted in North Shewa Zone of the Amhara region, Ethiopia; North Shewa Zone is one of the eleven Zones in Amhara Region. Prominent. North Shewa Zone subdivided into 24 districts. According to the 2007 national census conducted by the Central Statistical Agency of Ethiopia (CSA), North Shewa Zone had a total population of 1,837,490 of whom 928,694 are men and 908,796 women(18). In North Shewa Zone, it is estimated that around 2,558 mothers give births in each month. This study was conducted over the period of January 1-30/2023.

### Eligibility Criteria

All lactating mothers in North Shewa Zone with children aged 6-23 months during the study period were considered the source population. Mothers who had lived in the selected districts for at least six months formed the study population. Mothers with hearing impairments and physical deformities that prevented accurate anthropometric measurements were excluded from the study.

### Sample Size Determination and Sampling Techniques

The sample size was determined by considering a factor (age at first pregnancy: underweight for first pregnancy <18 years = 37.9%, underweight for first pregnancy ≥18 years = 23.1%)(19) which was significantly associated with outcome variable. The calculation was done using 95% confidence level, a margin of error 5%, and power of 80%, Using an open Epi info software program. By applying a design effect of 2 and adding a 10% non-response rate, the final sample size for this study was 722 lactating mothers.

The study participants were selected using a multistage sampling technique, assuming a uniform distribution of characteristics across all districts in the Zone. The Zone has 24 Districts and seven of them were randomly selected using a lottery method. All households with lactating mothers were identified from the health extension workers’ family folders by using simple random sampling. Households with at least one lactating mother were considered the sampling frame, and the lactating mother within the household was the sampling unit. If more than one lactating mothers were present in a household, one was selected using a lottery method. Up to three visits were made to each participant in case of absence during the initial visit.

### Data collection techniques and study variables

Data on the study variables were collected using semi-structured, pretested questionnaires in Amharic language, adapted from various literature sources(19–23). The adapted data collection tools, initially prepared in English, were translated into Amharic and then back-translated into English to ensure consistency.

The level of household food insecurity was measured using the Household Food Insecurity Access Scale (HFIAS), a structured, standardized and validated tool developed by FANTA. The mother’s dietary intake pattern was assessed through a qualitative recall of all food types consumed during the previous 24 hours. Then food items were categorized into food groups to calculated individual or women dietary diversity scores. The mean dietary diversity score was then used to classify maternal dietary intake as adequate or inadequate. Special food consumption days were excluded from the assessment.

To measure the outcome variable, anthropometric measurements were taken. The weight of each lactating mother was measured to the nearest 100 g using a portable electronic digital scale (Seca, Germany model), and height was measured to the nearest 0.1 cm using a portable wooden height measuring board with sliding head bar, following standard anthropometric measurement techniques.

### Data collectors and data collection procedures

The data were collected by seven diploma-holding data collectors residing in the study area. The data collection tool included questions related to socio-demographic variables, obstetric history, nutrition, health-related factors, and anthropometric measurements. Data collectors were trained for two days by the principal investigator.

Additionally, two BSc holders in Nursing were trained to supervise data collectors. Data were collected through home-to-home visit. Nutritional status of mothers was determined by using their BMI, calculated by dividing weight in kilograms by the square of height in meters. Based on the BMI values, mothers were categorized as underweight: less than 18.5 kg/m2, normal: 18.5-24.9 kg/m2, overweight: 25-29.9 kg/m2 and obese: ≥30 kg/m2 (28).

### Operational definition

Underweight: a woman who has a body mass index below 18.5 kg/m2 (24).

Feeding practice: eating habits of women during lactation(20).

Dietary diversity: the number of food groups that women consume during lactation(20).

Lactating mother: a woman who breastfeeds her child or infant during data collection(20).

### Data quality, processing and Analysis

Data quality was ensured through the careful design of the Semi-structured, pre-tested questionnaire. To minimize intra-observer variability, the relative technical error of measurement was calculated. Measurement results were standardized with those of the trainer during both the training and pretesting phases. Each data collector measured the height and weight of lactating mothers twice and recorded the average of the two measurements. The proper functioning of digital weight scales was checked before each use to ensure accuracy. A portable standard weight of 1 kg weight was used to verify the calibration of the scales. To further ensure data accuracy, double data entry was used to identify and resolve discrepancies.

The data entered into Epi-Data software were exported to SPSS version 25 software for further analysis. Descriptive statistics such as frequency tables, cross tabulations, prevalence, and graphical presentations were used. Binary and multivariable logistic regression analysis were performed to identify factors. Predicators with a p-value of less than 0.20 in the bivariate logistic regression were considered candidate variables for a multivariate logistic regression analysis, to control for potential confounders. Model fitness was check using the Hosmer-Lemeshow test and multi-collinearity was assessed before proceeding to multivariable analysis. In multivariable analysis, significant associations between variables and underweight were determined using the adjusted odds ratio (AOR) with a 95% confidence interval. Variables with a p-value less than 0.05 in the multivariable analysis were considered statistically significant.

## Result

### Socio-demographic characteristics

Out of the 722 sampled pregnant women, 678 responded to the questionnaires, resulting in a response rate of 93.9%. Various items were developed to assess the nutritional status of lactating mothers, as well as the socio-demographic determinant factors in the study area.

The mean age (± SD) of the participants was 26.3 (± 4.3) years, while the age range was 19-43 years. More than half of the respondents, 387(57.1%) were in the age range of 25-34 years. Most study respondents were married (94%).

As shown in Table 1, almost all respondents, 667(98.4%) were followers of orthodox religion followed by Muslims, 6(0.9%). Regarding ethnicity, the majority were Amhara, 657(96.9%) followed by Oromo, 15(2.2%).

**Table 1:**
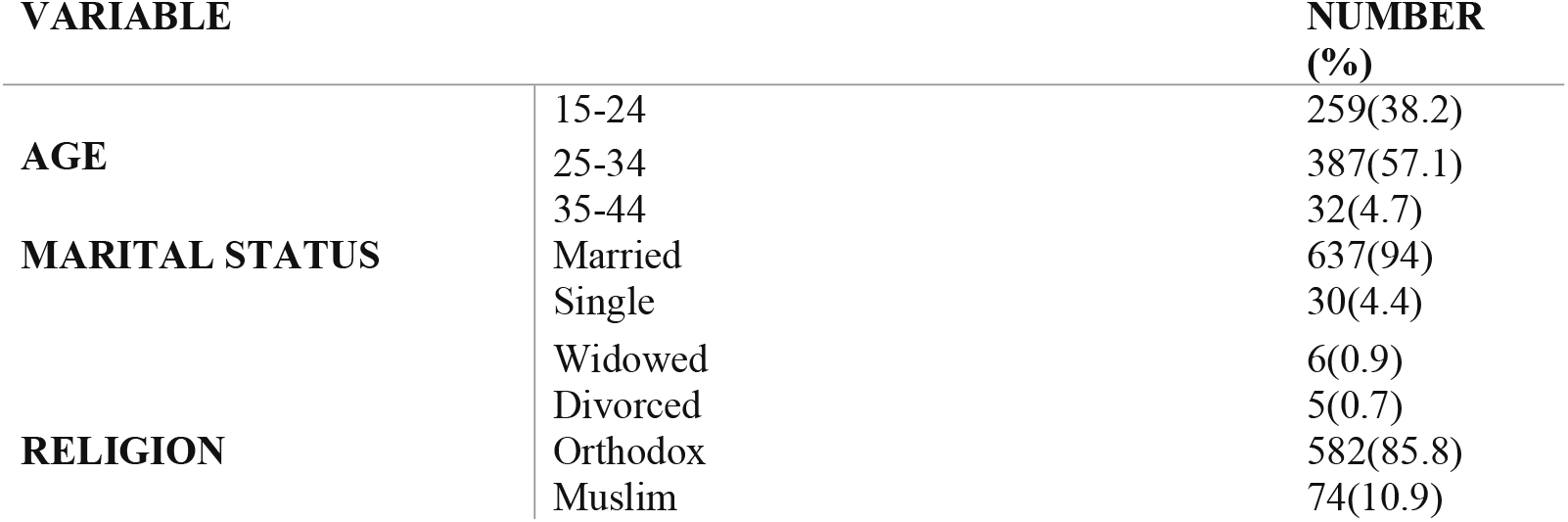
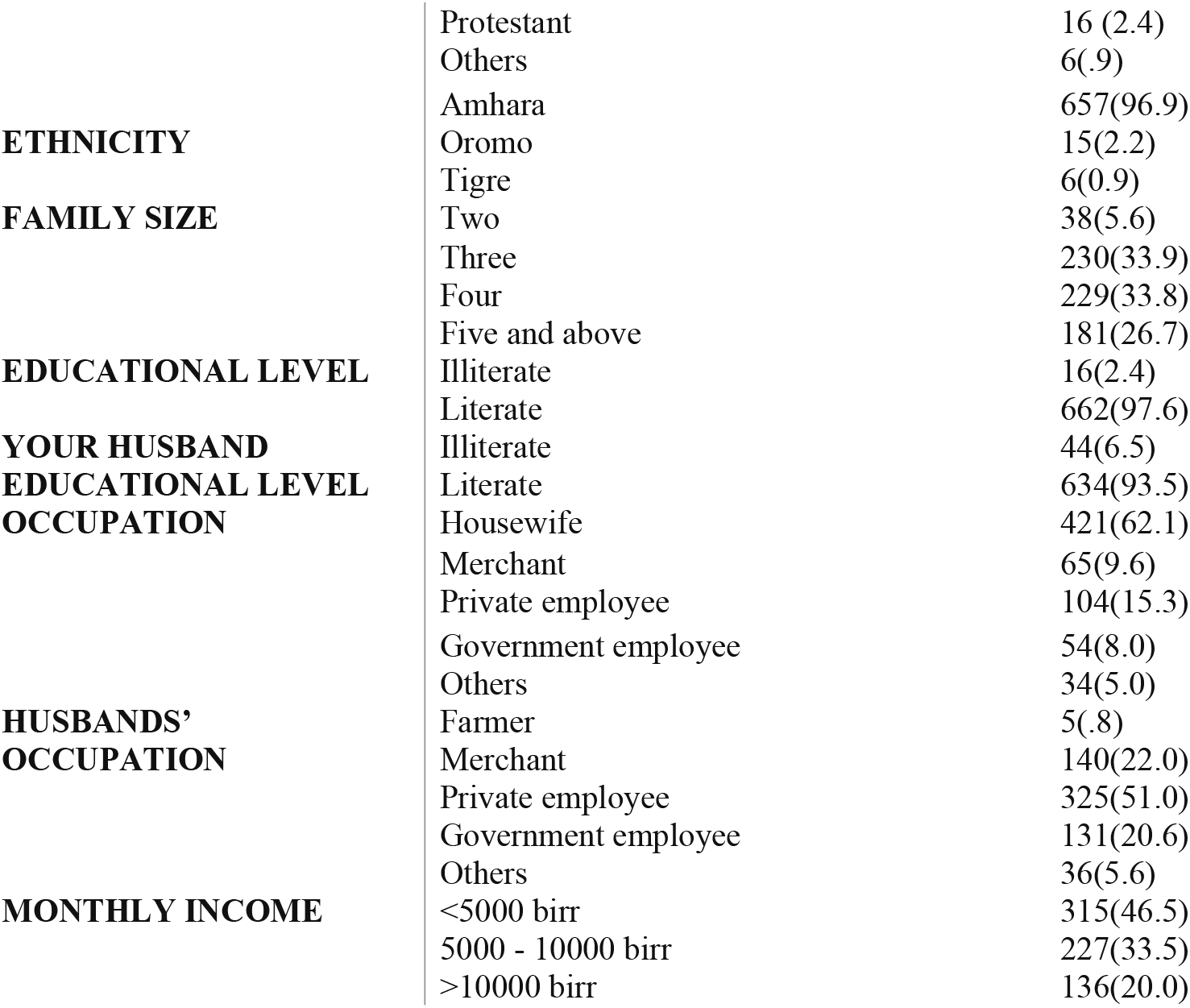
Distribution of Socio-demographics characteristics of lactating mothers in North Shewa Zone, Amhara region, Ethiopia, 2023 (N= 678).

In terms of family size, 30.7% and 26.4% of mothers had three and two family members, respectively.

Of the 678 respondents, about one third, 228(33.6%) had either completed primary school or had no formal education, while the remaining had attained secondary school education or higher.

Regarding the husbands’ educational level, about two thirds, 409(64.2) had education beyond primary school. The majority of respondents, 421(62.1%) were homemakers and about half of the husbands, 325(47.9 %) were employed in private organizations.

### Medical and obstetric Characteristics

Table 2 depicts medical and obstetric characteristics of lactating mothers. About two-thirds, 458(67.6%) of lactating mothers had four or more antenatal care visits, and around half, 346(51%) had at least one postnatal follow-up during their most recent lactation period.

**Table 2:**
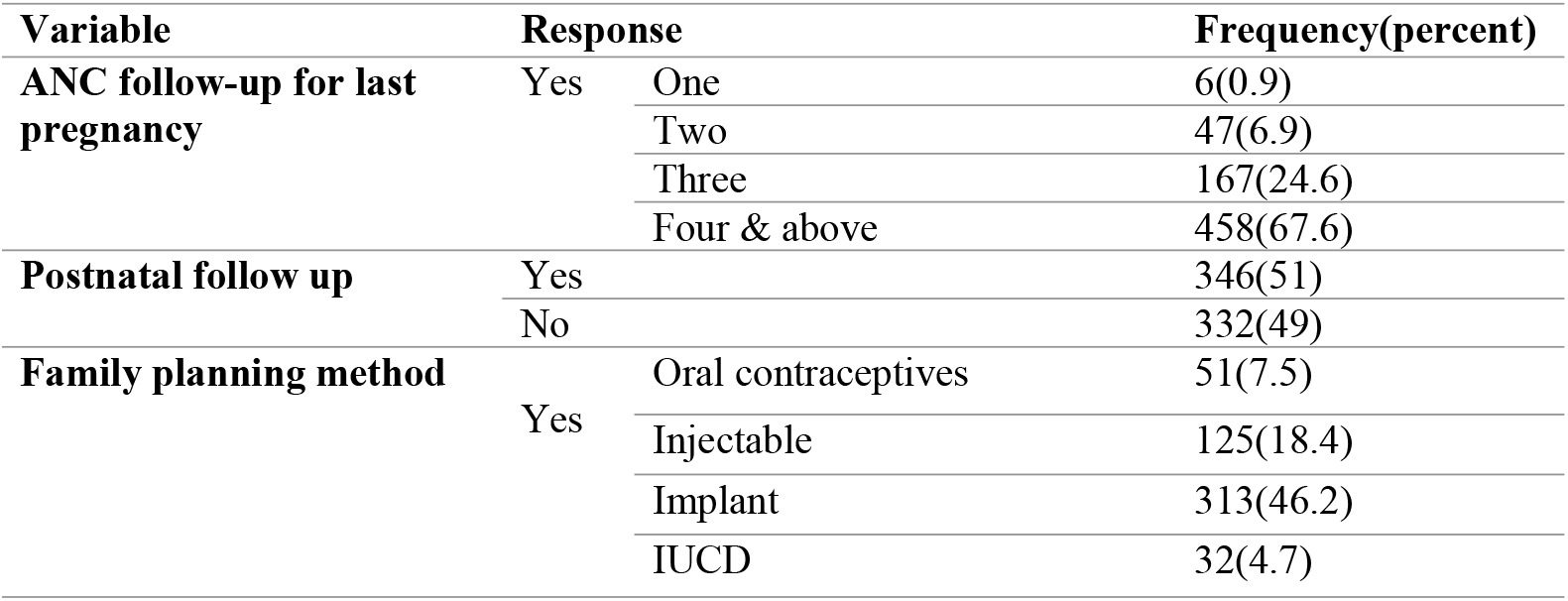
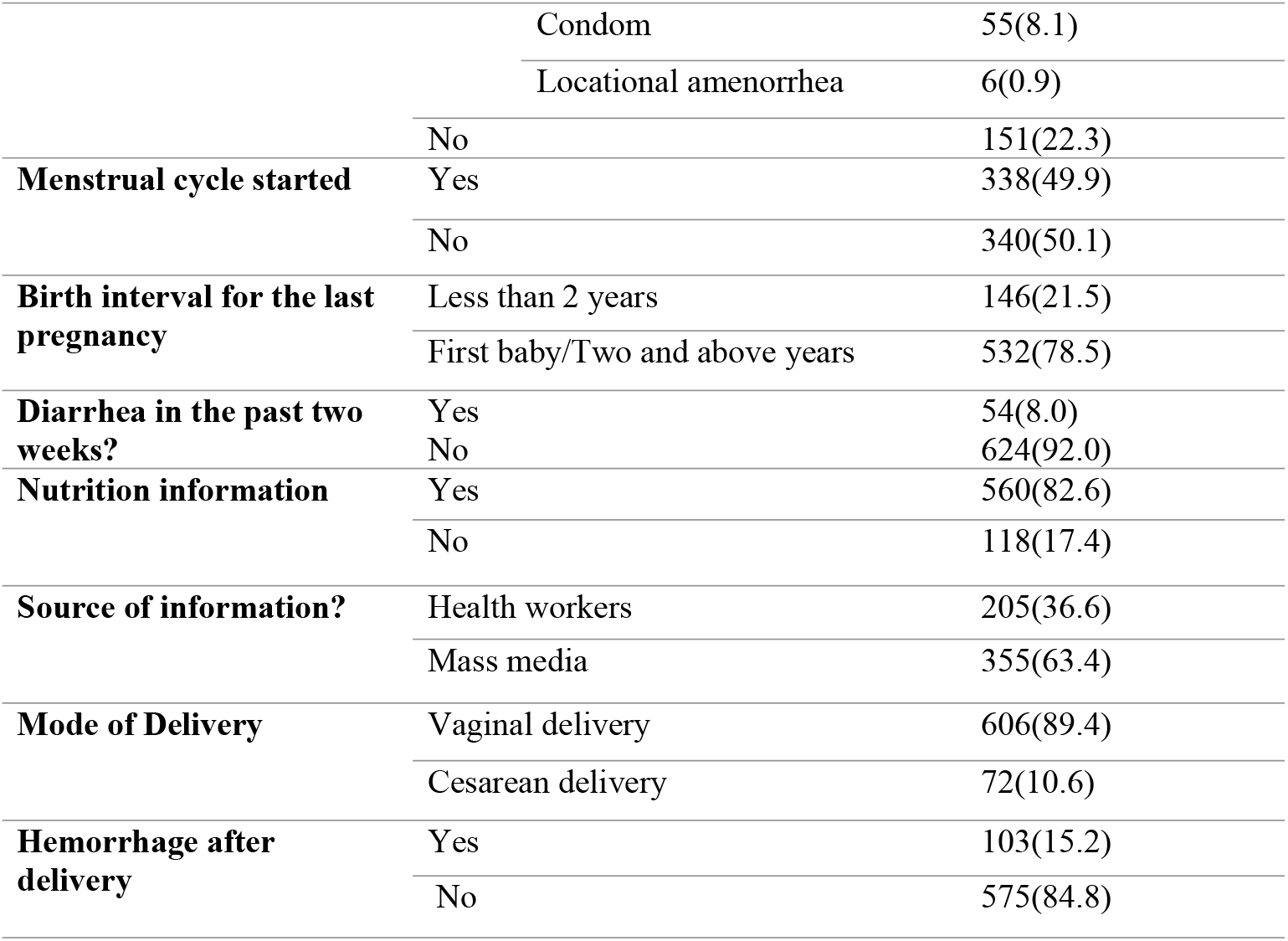
Distribution of Obstetrics and medical characteristics of lactating mothers in North Shewa Zone, Amhara region, Ethiopia, 2023 (N= 678).

| Variable | Response |  | Frequency(percent) |
| --- | --- | --- | --- |
| **ANC follow-up for last pregnancy** | Yes | One | 6(0.9) |
|  |  | Two | 47(6.9) |
|  |  | Three | 167(24.6) |
|  |  | Four & above | 458(67.6) |
| **Postnatal follow up** | Yes |  | 346(51) |
|  | No |  | 332(49) |
| **Family planning method** | Yes | Oral contraceptives | 51(7.5) |
|  |  | Injectable | 125(18.4) |
|  |  | Implant | 313(46.2) |
|  |  | IUCD | 32(4.7) |

|  |  |  |
| --- | --- | --- |
|  | Condom | 55(8.1) |
|  | Locational amenorrhea | 6(0.9) |
|  | No | 151(22.3) |
| **Menstrual cycle started** | Yes | 338(49.9) |
|  | No | 340(50.1) |
| **Birth interval for the last pregnancy** | Less than 2 years | 146(21.5) |
|  | First baby/Two and above years | 532(78.5) |
| **Diarrhea in the past two weeks?** | Yes | 54(8.0) |
|  | No | 624(92.0) |
| **Nutrition information** | Yes | 560(82.6) |
|  | No | 118(17.4) |
| **Source of information?** | Health workers | 205(36.6) |
|  | Mass media | 355(63.4) |
| **Mode of Delivery** | Vaginal delivery | 606(89.4) |
|  | Cesarean delivery | 72(10.6) |
| **Hemorrhage after delivery** | Yes | 103(15.2) |
|  | No | 575(84.8) |

The majority of mothers, 522(77%) had used contraceptives to prevent pregnancy. About one tenth, 72(10.6%) gave birth via cesarean section, and approximately one-seventh, 103(15.2%) experienced postpartum hemorrhage.

### Meal Habits of the Household

About half of the respondents, 344 (50.7%) reported having four or more meals per day. About two-thirds, 441(65%) of mothers received the minimum acceptable diet per day. Approximately one-tenth, 64(9.4%) of lactating mothers experienced household food insecurity during the data collection period (see Table 3).

**Table 3:** Distribution of meal frequency, diet diversity and Household Food Security Status of Lactating Mothers in North Shewa Zone, Amhara Region, Ethiopia, 2023 (N= 678).

| **Variable** | **Status** | **Frequency (percent)** |
| --- | --- | --- |
| **Frequency of meal per day** | 2-3 | 334(49.3) |
|  | 4-5 | 344(50.7) |
| **Having the minimum acceptable diet** | Yes | 441(65) |
|  | No | 237(35) |
| **Household food security** | Secured | 614(90.4) |
|  | Insecure | 64(9.4) |

### Nutritional status of lactating mothers

Anthropometric measurements showed that the women had a mean BMI of 22.9 ±3.5 kg/m2. Among respondents, 42(6.3%), 126(18.7%), 58(8.6%) of the women are obese, over weight and underweight respectively (see Fig.1).

**Figure 1:**
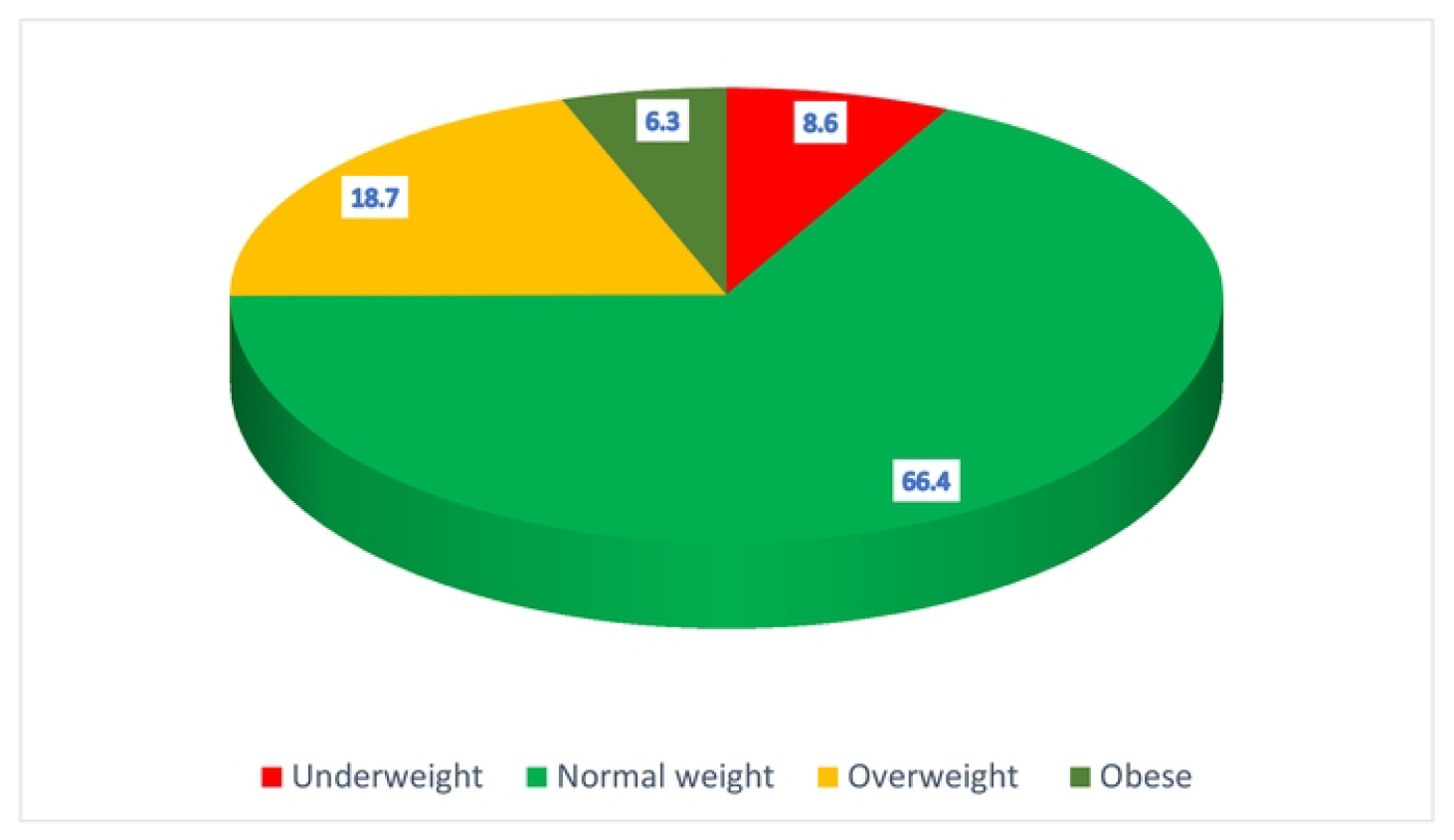
Distribution of Nutritional Status based on BMl Among Lactating Mothers in North Shewa Zone, Ethiopia, 2023. (n = 678)

### Bi-variable and Multivariable Analysis of Factors Associated with Undernutrition Among Lactating mothers

In this study, the bivariable analysis revealed educational status, occupation, family size, pregnancy interval, parity, duration of breastfeeding, Postnatal follow up, return of menstrual cycle, heavy bleeding after delivery and contraceptives use were statistically associated with the undernutrition (<18.5Kg/m^2)^ among mothers (p<0.02).

After bivariable analysis, predictors with a p-value less than 0.2 were included in the multivariable analysis. In a multivariable analysis: housewives were 4 times more likely to be undernourished compared to mothers working outside the home (AOR=4, 95% CI: 2.8,8.2), Mothers from households with family size of five or more were about 4 times more likely to be undernourished than those with fewer than 5 family members (AOR=3.8, 95% CI: 1.1,12.6). Mothers who breastfed for more than 12 months were about 4 times more likely to be undernourished than those who breastfed for 12 months or less (AOR=4.2, 95% CI: 2.6,7.9), Mothers who had no postnatal follow up were 6 times more likely to be undernourished than those who received postnatal care (AOR=6, 95% CI: 2.3,11.9), and respondents who had not started using contraceptives were 5 times more likely to be undernourished than who had begun using contraceptives (AOR=5.1, 95% CI: (2.2,12.6) as shown in Table 4.

**Table 4:**
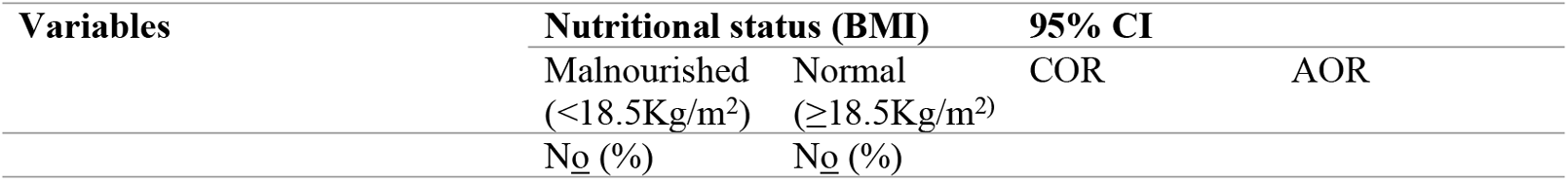
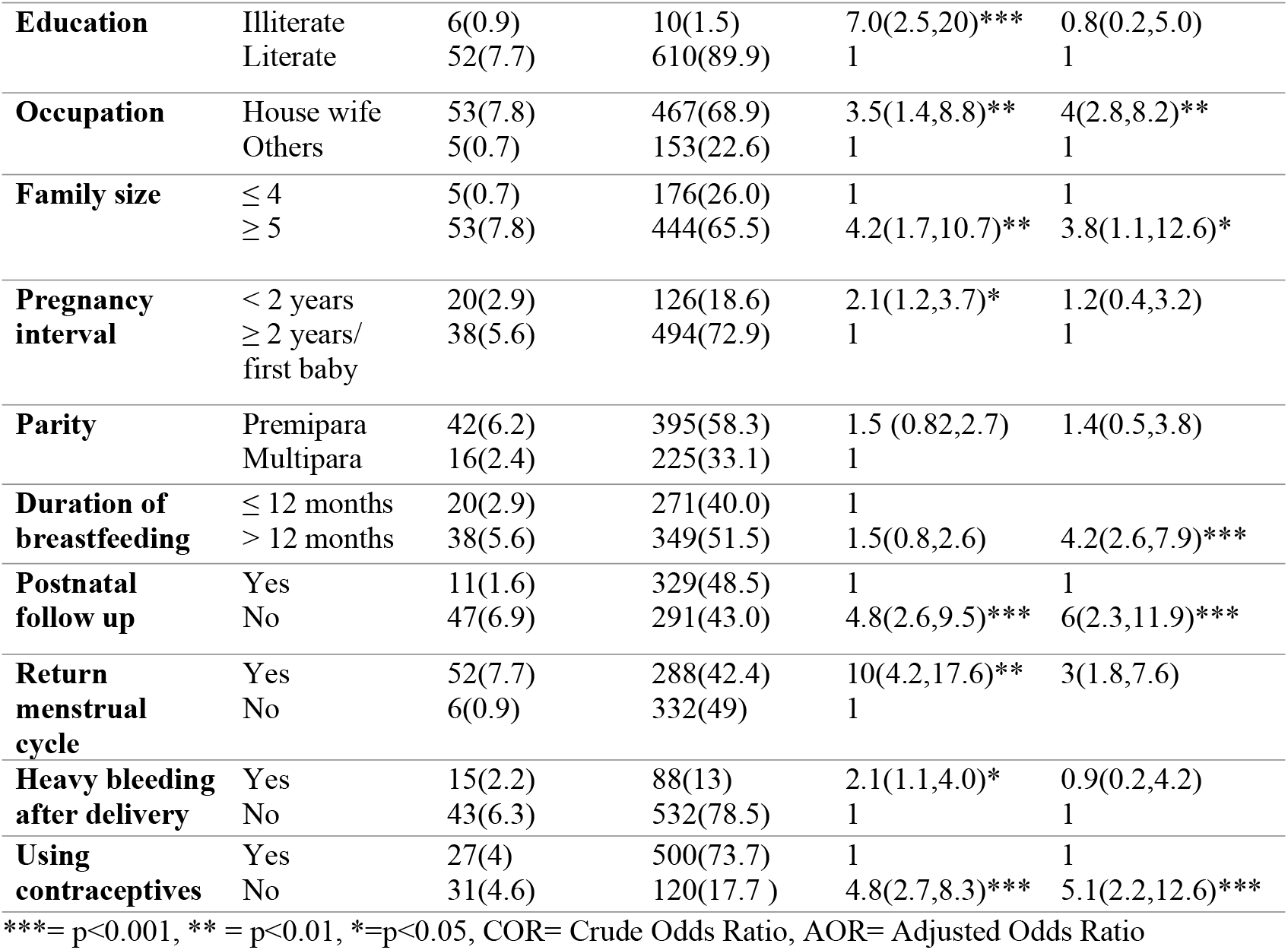
Variables with the Underweight base on (BMI) of Lactating Mothers in North Shewa Zone, Ethiopia, 2023 (n = 678)

| **Variables** | **Nutritional status (BMI)** |  | **95% CI** |  |
| --- | --- | --- | --- | --- |
|  | Malnourished (<18.5Kg/m<sup>2</sup>) | Normal (≥18.5Kg/m<sup>2</sup>) | COR | AOR |
|  | No (%) | No (%) |  |  |

|  |  |  |  |  |  |
| --- | --- | --- | --- | --- | --- |
| **Education** | Illiterate | 6(0.9) | 10(1.5) | 7.0(2.5,20)*** | 0.8(0.2,5.0) |
|  | Literate | 52(7.7) | 610(89.9) | 1 | 1 |
| **Occupation** | House wife | 53(7.8) | 467(68.9) | 3.5(1.4,8.8)** | 4(2.8,8.2)** |
|  | Others | 5(0.7) | 153(22.6) | 1 | 1 |
| **Family size** | ≤ 4 | 5(0.7) | 176(26.0) | 1 | 1 |
|  | ≥ 5 | 53(7.8) | 444(65.5) | 4.2(1.7,10.7)** | 3.8(1.1,12.6)* |
| **Pregnancy interval** | < 2 years | 20(2.9) | 126(18.6) | 2.1(1.2,3.7)* | 1.2(0.4,3.2) |
|  | ≥ 2 years/<br>first baby | 38(5.6) | 494(72.9) | 1 | 1 |
| **Parity** | Premipara | 42(6.2) | 395(58.3) | 1.5 (0.82,2.7) | 1.4(0.5,3.8) |
|  | Multipara | 16(2.4) | 225(33.1) | 1 |  |
| **Duration of breastfeeding** | ≤ 12 months | 20(2.9) | 271(40.0) | 1 |  |
|  | > 12 months | 38(5.6) | 349(51.5) | 1.5(0.8,2.6) | 4.2(2.6,7.9)*** |
| **Postnatal follow up** | Yes | 11(1.6) | 329(48.5) | 1 | 1 |
|  | No | 47(6.9) | 291(43.0) | 4.8(2.6,9.5)*** | 6(2.3,11.9)*** |
| **Return menstrual cycle** | Yes | 52(7.7) | 288(42.4) | 10(4.2,17.6)** | 3(1.8,7.6) |
|  | No | 6(0.9) | 332(49) | 1 |  |
| **Heavy bleeding after delivery** | Yes | 15(2.2) | 88(13) | 2.1(1.1,4.0)* | 0.9(0.2,4.2) |
|  | No | 43(6.3) | 532(78.5) | 1 | 1 |
| **Using contraceptives** | Yes | 27(4) | 500(73.7) | 1 | 1 |
|  | No | 31(4.6) | 120(17.7) | 4.8(2.7,8.3)*** | 5.1(2.2,12.6)*** |

\*\*\*= p<0.001, \*\* = p<0.01, \*=p<0.05, COR= Crude Odds Ratio, AOR= Adjusted Odds Ratio

## Discussion

The aim of this study was to investigate the nutritional status and associated factors among lactating women in North Shewa zone, Amhara Region, Ethiopia, using Body Mass Index (BMI) based on WHO standard classification. Most mothers, 450(66.4%) had a normal BMI (18.5 kg/m^2^ - 24.9 kg/m^2^), while 58(8.6%) were underweight, 126(18.7%) overweight, and 42(6.3%) obese.

This reflects the growing “double burden” of malnutrition: coexistence of under and over-nutrition in the same population. The combined over-weight/obese prevalence (25%) signals an emergent public health challenge. Overnutrition have implications not just for maternal health such as hypertension, diabetes, and complications in lactation, but also for child outcomes, including increased risk of obstetric complications and potential long-term metabolic effects in offspring (25).

A similar community-based cross-sectional study conducted in rural Tibetan using anthropometric measurement (BMI) showed that 10.3% of a mother had a body mass index of less than 18.5 kg/m^2^ (26). The underweight prevalence in this study (8.6%) is notably lower than that reported in Yilmana Densa district, Ethiopia, where 22.6% (3), in Dessie town about 21%(27), in Sekota camps 25%(4) of lactating mothers were undernourished. It is dramatically lower than the 55.2% prevalence observed in war affected Tigray region(8). The comparatively better outcomes in North Shewa may reflect improved access to postnatal care, nutrition counseling and community level maternal initiatives. In contrast conflict affected zone such as in Tigray reported impaired healthcare infrastructure and disrupted food supply chains that exacerbated undernutrition.

### Associated Factors of Undernutrition Among Lactating Mothers

Lactating mothers who worked as housewives were significantly more likely to be undernourished compared to women employed outside the home. A community-based study in Nekemte, Oromia, reported similar findings, revealing that housewives had elevated odds of undernutrition (28), likely due to lower education levels, limited decision-making autonomy, and reduced financial control.

Mothers from households with five and more members faced a high risk of being undernourished. Studies from Nekemte, and Chiro, Ethiopia and Accra, Ghana, similarly showed that large household sizes were associated with maternal undernutrition. This may be due to increased caregiving demands and shared limited resources, stretching both time and food availability for the mothers (15, 28, 29).

Extended breastfeeding, particularly beyond 12 months, was associated with higher odds of undernutrition. The Tigray study found that mothers breastfeeding beyond one year had 2.8 times higher odds of being underweight, suggesting cumulative nutrient depletion without adequate dietary compensation (20, 30).

Mother who did not attend postnatal care visits were significantly associated with undernutrition up to sixfold higher odds. Similar study from Yimana densa district, Ethiopia, showed that postnatal follow up had an effect on nutritional status of lactating mothers (3). Postnatal care provides vital nutritional advice, supplementation, and health screening, all of which can protect maternal nutritional status after delivery.

The last predictor variable in this study was contraceptive utilization. Similar study from Nekemte, Ethiopia, showed that non-use of modern contraceptives increased the risk of undernutrition, with mothers who had not begun contraception more likely to be underweight. Proper birth spacing through contraceptive use allows the mothers’ body time to replenish nutritional reserves between pregnancies (28).

Generally, this study was conducted with considering to minimize study limitations, the data collectors failed to reach the remote areas in North Shewa Zone and the investigators managed them as non-respondents. Recall bias and lack of micronutrient data may also underestimate certain factors.

## Conclusion

The findings revealed a double burden of malnutrition while two-thirds of lactating mothers in North Shewa Zone were in normal weight range. The findings of this study showed the magnitude of underweight lactating mothers was relatively low when comparing with similar communities in Ethiopia.

Undernutrition among lactating mothers was associated with several socio-demographic and reproductive factors, including being housewife, large family size, extended breastfeeding beyond 12 months, lack of postnatal care and non-use of contraceptives. These findings underscore the importance of strengthening maternal health programs that support postnatal follow-up, nutritional counselling, family planning and economic empowerment particularly for housewives.

## Abbreviations

ANC: Antenatal Care
AOR: Adjusted Odds Ratio
BMI: Body Mass Index
CED: Chronic Energy Deficiency
CSA: Central Statistics Agency
DDS: Dietary Diversity Score
FAO: Food and Agricultural Organization
GNR: Global Nutrition Report
HFIAS: Household Food Insecurity Access Scale
IFAD: International Fund for Agricultural Development
MUAC: Mid Upper Arm Circumference
NGO: Non-Governmental Organization
PNC: Post-natal Care
SPSS: Statistical Package for Social Science
UNICEF: United Nation International Children Emergency Fund
UNSCN: United Nation Standing Committee for Nutrition
WFP: World Food Program
WHO: World Health Organization

## Ethical consideration

This study was reviewed and approved by the IRB of Asrat Woldeyes Health Science Campus, Debre Berhan University and official letter was sent to North Shewa Zone health office and data collection began after permission was granted and in cooperation with the selected districts. prior to data collection, written informed consent was obtained from all participants after explaining the aim, procedures, potential risk factors and benefits of the study in local language (Amharic). participants were also assured of the confidentiality of their responses and their right to withdraw from the study at any point without any consequences.

## Availability of the data statement

The data will be available from the corresponding author on reasonable request

